# Alterations in pathogen-specific cellular and humoral immunity associated with acute peripheral facial palsy of infectious origin

**DOI:** 10.1101/2023.08.13.23294046

**Authors:** Leyla Mohammad, Mathias Fousse, Gentiana Wenzel, Marina Flotats-Bastardas, Klaus Faßbender, Ulrich Dillmann, Bernhard Schick, Michael Zemlin, Barbara C. Gärtner, Urban Sester, David Schub, Tina Schmidt, Martina Sester

## Abstract

**Background and Objectives:** Peripheral facial palsy (PFP) is a common neurologic symptom which can be triggered by pathogens or autoimmunity as well as trauma, tumors, cholesteatoma and further local conditions disturbing the peripheral section of the nerve. In general, its cause is often difficult to identify, remaining unknown in over two thirds of cases. As we have previously shown that the quantity and quality of pathogen-specific T cells change during active infections, we hypothesized that such changes also may help to identify the causative pathogen in PFPs of unknown origin.

**Methods:** Pathogen-specific T cells were quantified in blood samples of 55 patients with PFP and 23 healthy controls after stimulation with antigens from varicella-zoster virus (VZV), herpes-simplex viruses (HSV) or borrelia. T cells were further characterized by expression of the inhibitory surface molecule CTLA-4, and markers for differentiation (CD27) and proliferation (Ki67). Pathogen-specific antibody responses were analyzed using ELISA. Results were compared with conventional diagnostics.

**Results:** Patients with PFP were more often HSV-seropositive than controls (p=0.0003), whereas VZV-and borrelia-specific antibodies did not differ between groups. Although the quantity and general phenotypical characteristics of antigen-specific T cells did not differ either, expression of CTLA-4 and Ki67 was highly increased in VZV-specific T cells of 9 PFP patients, of which 5 showed typical signs of cutaneous zoster. In the remaining 4 patients, a causal relationship with VZV was possible but remained unclear by clinical standard diagnostics. A similar CTLA-4- and Ki67-expression profile was also found in a patient with acute neuroborreliosis.

**Discussion:** In conclusion, the high prevalence of HSV-seropositivity among PFP-patients may indicate an underestimation of HSV-involvement in PFP, even though HSV-specific T-cell characteristics seem insufficient to identify HSV as a causative agent. In contrast, striking alterations in VZV- and borrelia-specific T-cell phenotype and function may allow identification of VZV- and borrelia-triggered PFPs thus bearing the potential to improve specificity of the clinical diagnosis.

## Introduction

Peripheral facial palsy (PFP) is a common neurological symptom consisting of an incomplete or complete loss of signal transmission by the facial nerve resulting in a variable degree of mainly unilateral palsy of mimic muscles. The severity of symptoms is indicated by House-Brackmann grading (1). Approximately 60-75% of patients are diagnosed with idiopathic PFPs of unknown origin, also termed Bell’s palsy (2). Herpes-simplex virus (HSV) has been suggested to be causally related to idiopathic PFPs to some extent, although its involvement is still controversially discussed (3, 4). In general, non-idiopathic PFPs are often triggered by infectious pathogens, with most frequent involvement of varicella-zoster virus (VZV; with or without simultaneous skin disease), or borrelia (neuroborreliosis) (5-9). In addition, although less frequent, administration of vaccines, or autoimmune or neoplastic diseases can also be causally linked to acute PFPs, and trauma, tumors, cholesteatoma and further local conditions disturbing the function of the peripheral part of the nerve have as well to be ruled out in the first stage of diagnoses (10-12).

The choice of a specific therapeutic regimen is difficult, especially for patients with idiopathic PFP, where sufficiently powered studies are lacking (2, 13). As a consequence, depending on provisional evaluation during the first visit, PFPs are usually treated with steroids, anti-viral and/or anti-bacterial agents. Patients are treated with single agents directed against the most probable cause, although an inappropriate choice of treatment may bear the risk of prolonged symptoms or permanent damages of the nerve (13, 14). Current evidence recommends a combination therapy in uncertain cases or when pathogen diagnosis is delayed (15, 16). Therefore, a faster and more specific diagnosis of the causative agent of PFP would facilitate a more directed and specific choice of therapy.

Cerebral imaging may be considered for additional symptoms to detect neoplastic processes or brainstem lesions (17). Electrophysiological procedures provide evidence of early hypoexcitability in the facial canal (typical in idiopathic PFP), but are not specific for the etiology of PFP (18, 19). Although idiopathic PFP is typically characterized by absence of detectable pathogens in the cerebrospinal fluid (CSF), analysis of CSF is useful for determination of highly increased numbers of immune cells (pleocytosis), of pathogenic nucleic acids, and/or of intrathecal increase of pathogen-specific antibodies compared to blood (antibody specific index, ASI) (20-22). However, a lumbar puncture is sometimes not possible due to limited compliance, anatomical abnormalities or coagulations disorders (23). In general, less invasive detection of pathogenic nucleic acids and pathogen-specific antibodies (IgA or IgM) from blood samples may be of value, but the diagnostic windows for nucleic acid detection in blood is comparatively small and specificity of positive IgA and IgM status as marker for active involvement of the respective pathogen is limited. Therefore, based on the limited availability of specific diagnostic approaches, identification of the underlying cause of PFP is difficult thereby resulting in non-specific or symptomatic treatment regimens.

In recent years, the analysis of pathogen-specific T-cell responses has shown that effective pathogen control is not only determined by the quantity of pathogen-specific T cells but also depends on their functional and phenotypic properties. For instance, active infection with cytomegalovirus (CMV) after solid organ transplantation was associated with an increase in the expression of the inhibitory surface molecule PD-1 on CMV-specific T cells (24). Likewise, we have previously shown that VZV-specific T cells during active herpes zoster in immunocompetent and immunocompromised individuals show alterations in phenotype characterized by an increased expression of CTLA-4 and PD-1 (25). These VZV-specific T-cell characteristics are distinct from those of non-symptomatic individuals (25) and were not only found in patients with skin rashes, but also in patients with VZV-related central nervous system (CNS) infections (26). Thus, an increased CTLA-4 expression may serve as a highly sensitive marker for identification of VZV-related CNS-infections such as meningitis or encephalitis, even in the absence of a VZV-rash (26). We therefore hypothesized that alterations in pathogen-specific T cells may be used to identify the causative pathogen in patients with PFPs.

## Results

### Study population

Fifty-five patients with PFP and 23 healthy controls were recruited. Demographic and clinical characteristics of the study population are summarized in table 1. Patients showed higher leukocyte counts (p=0.042), and their percentage of lymphocytes was significantly lower (p=0.002). The neutrophil/lymphocyte ratio as a prognostic hematologic marker of Bell’s Palsy (27) was also significantly higher in patients than in controls (p=0.004). The intensity of PFP indicated by the House-Brackmann grading was moderate to high. Based on results of standard clinical parameters and procedures, 32 PFP-patients were treated with steroids only and one with an anti-bacterial agent, whereas 17 received a combination of steroids, anti-viral and/or anti-bacterial agents as double (n=9) or triple drug regimen (n=8). Two patients received intravenous immunoglobulins only based on suspected Guillain-Barré syndrome. Three patients received symptomatic treatment only. CSF was obtained in 44/55 patients (80%), whereas lumbar puncture was not possible in 11 cases due to either oral anticoagulation (n=6) or lack of consent (n=5).

**Table 1:** Characteristics of the study populations.

|  | PFP-patients | controls | p-value |
| --- | --- | --- | --- |
| n | 55 | 23 |  |
| Years of age [mean±SD] | 46.89±19.3 | 43.1±17.4 | 0.413 |
| Females [n (%)] | 21 (38.2%) | 14 (60.9%) | 0.089 |
| Leukocytes/μl blood<br>[median (IQR)] | 7200 (3260) | 6980 (2370) | 0.042 |
| % lymphocytes in blood<br>[median (IQR)] | 26.0 (11.1) | 33.1 (5.4) | 0.002 |
| Neutrophil/lymphocyte ratio<br>[median (IQR)] | 2.42 (1.47) | 1.73 (0.48) | 0.004 |
| Days between 1<sup>st</sup> and 2<sup>nd</sup><br>blood sampling, [mean±SD] | 13.8±2.1<br>(n=36) | 14.0±1.0<br>(n=15) | 0.996 |
| Clinical characteristics PFP |  |  |  |
| days since onset of symptoms<br>[mean±SD] | 2.6±2.1 | n.a. |  |
| House-Brackmann grading (1-6)<br>[median (IQR)]<sup>#</sup> | 3.5 (1.5) | n.a. |  |
| Lumbar puncture | (n=44) | n.a. |  |
| normal cell counts (0-4 cells/μl) | 32 (72.7%) | n.a. |  |
| pleocytosis (≥5 cells/μl)<sup>*</sup> | 12 (27.3%) | n.a. |  |
| PFP-treatment [n]<sup>§</sup> | 52 | n.a. |  |
| steroids | 32 |  |  |
| anti-bacterial<sup>b</sup> | 1 |  |  |
| steroids/anti-viral<sup>a</sup> | 2 |  |  |
| anti-viral<sup>a</sup>/anti-bacterial<sup>b</sup> | 7 |  |  |
| steroids/anti-viral<sup>a</sup>/anti-bacterial<sup>b</sup> | 8 |  |  |
| immunoglobulins | 2 |  |  |

<sup>#</sup>House-Brackmann grading was not available in 3 patients; <sup>§</sup>3 patients with symptomatic treatment only; <sup>\*</sup>median 14 (IQR 70.8) cells/μl; <sup>a</sup>i.e. (val)aciclovir, <sup>b</sup>i.e. ceftriaxone, doxycycline or ampicillin.

### Correlation of pathogen-specific T-cell frequencies and antibody levels in patients and controls

As VZV, HSV and borrelia are among the main causes of infectious PFP, specific humoral and cellular immune responses against these pathogens were analyzed. In addition, CMV-specific immune responses were assessed as an internal control, as its involvement with PFP is rare. The serostatus for the respective pathogens among PFP-patients and controls is shown in table 2. In addition to IgG-levels, IgA-(VZV and HSV) and IgM-(HSV and borrelia) status were assessed. While most individuals were VZV-IgG positive, positive or intermediate VZV-IgA levels were found in 43.6% of PFP-patients and 21.7% of controls (p=0.078, table 2). Unlike for the other pathogens, the percentage of HSV-IgG positive individuals was significantly higher in the studied PFP-patients (90.9%) than in controls (52.2%, p=0.0003). Concerning other HSV Ig-classes, 47.3% of the tested individuals were IgA-positive, and 3.6% were IgM-positive with no difference between the groups (p=0.133 and p=0.470, respectively). Of note, the percentages of individuals with borrelia-IgG and IgM was low in both groups, and only one patient had both borrelia-IgG and IgM. As expected from German seroprevalence, about 50% of all individuals were CMV IgG-positive.

**Table 2:**
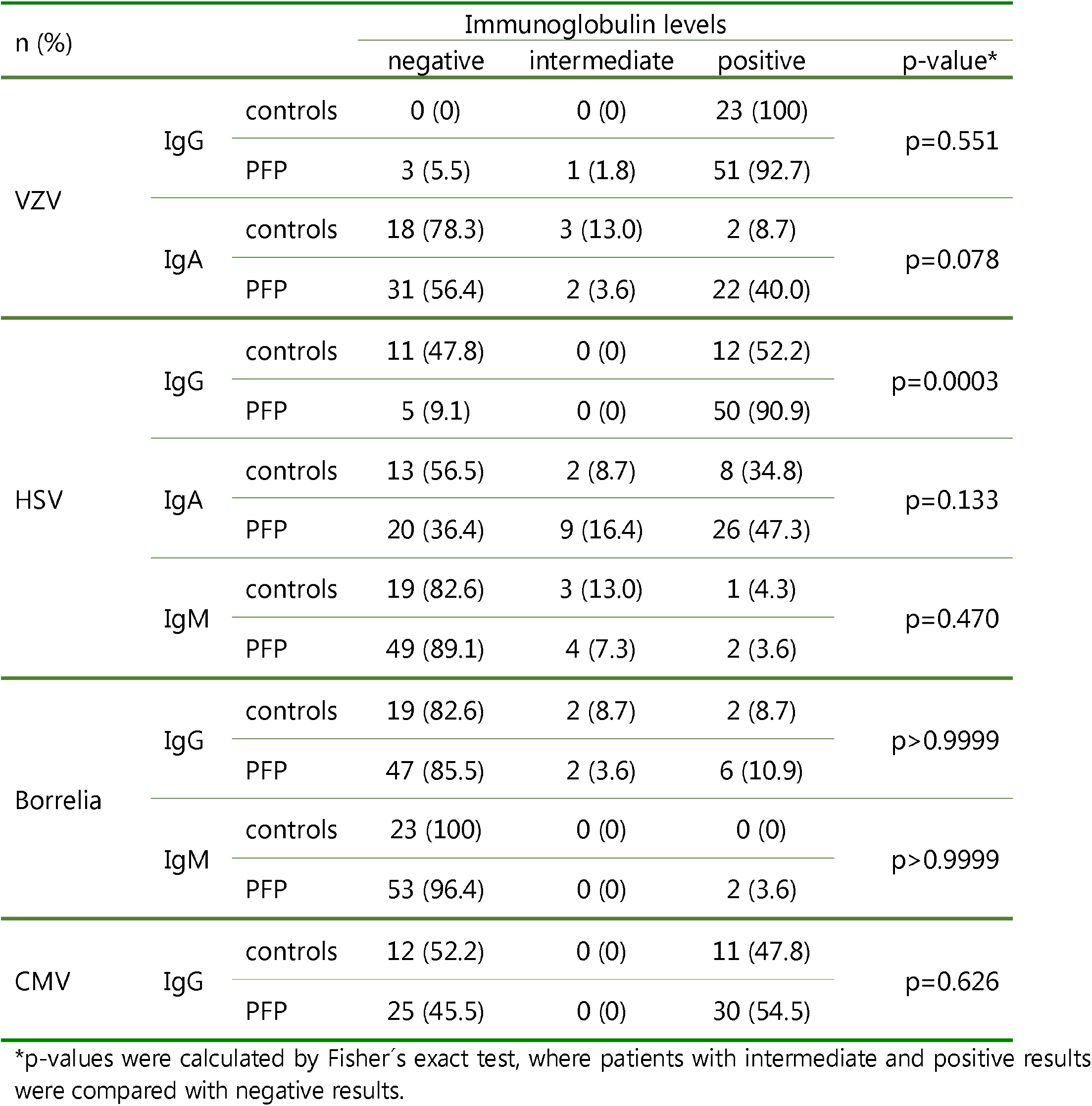
Serostatus of the study population.

To assess the quantity of the pathogen-specific T-cells, whole blood samples of controls and PFP-patients were stimulated with pathogen-derived antigens in vitro. Stimulation with SEB was carried out to quantify polyclonal T-cell responses. Representative dot plots of CD4 T cells of a 36-years old healthy control after stimulation with negative control antigen, VZV-antigen and SEB are shown in figure 1A. Median percentages of pathogen-specific T cells did not differ between PFP-patients and controls, independent of their serostatus (figure 1B and C, all p>0.05). Except for borrelia-specific immunity, T-cell frequencies were generally highest for IgG-seropositive individuals, and the percentage of pathogen-specific T-cells showed a significant correlation with corresponding IgG-levels (figure 1D).

**Figure 1:**
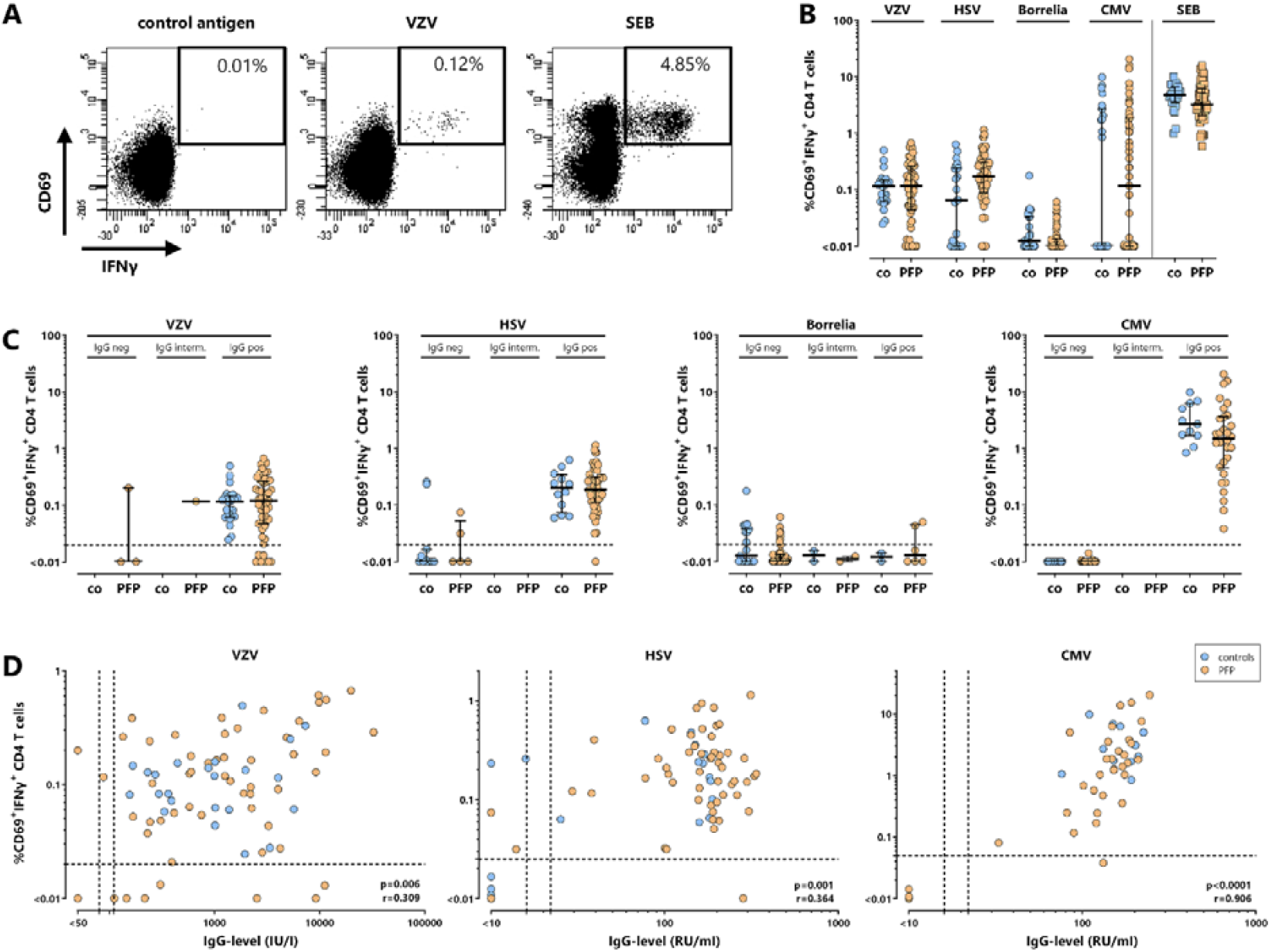
Correlation of pathogen-specific T cells and antibodies in patients with acute PFP and healthy controls. **(A)** Representative dot plots of CD4 T cells of a 36-years old healthy control are shown after stimulation with either control lysate (control), VZV-lysate (VZV) or polyclonal stimulus *Staphylococcus aureus* enterotoxin B (SEB, positive control). Numbers indicate percentages of reactive CD4 T cells of total CD4 T cells and are characterized by co-expression of the activation marker CD69 and the cytokine IFN*γ*. In **(B)** percentages of VZV-, HSV-, borrelia-, CMV-specific and SEB-reactive CD4 T cells were compared between 23 controls (co, blue circles) and 55 patients with PFP (orange circles). **(C)** Percentages of pathogen-specific T cells in controls and PFP-patients were stratified according to the corresponding IgG-serostatus, whereas in **(D)** these percentages were correlated with corresponding IgG-levels (VZV, HSV, CMV). As determination of borrelia-specific antibodies relied on a two-step screening and confirmation system with semi-quantitative output, analysis of borrelia-specific IgGs was restricted to semi-quantitative analysis. In panel B and C median values are indicated for each group. There are no significant differences between controls and PFP-patients in panels B and C. Dotted lines in panel C and D represent detection limits for pathogen-specific CD4 T cells or IgG-levels. CMV, cytomegalovirus, HSV, herpes-simplex viruses; IFN, interferon; PFP, peripheral facial palsy; VZV, varicella-zoster virus.

### No distinct differences in phenotypical and functional parameters of pathogen-specific T-cell responses between patients and controls

To analyze pathogen-specific cellular immunity in more detail, T cells from all patients and controls were phenotypically and functionally characterized based on expression of CTLA-4, CD27 and Ki67 (figure 2A). The analyses shown in figure 2 were restricted to all samples with detectable pathogen-specific T cells, and their percentage did not differ between the two groups (figure 2B). Moreover, median expression levels of the inhibitory receptor CTLA-4 were generally low (MFI<1000), despite high interindividual variabilities for VZV-, HSV- and CMV-specific T cells. Interestingly, although there were no significant differences between controls and PFP-patients, VZV-specific T cells of 9 patients showed clearly higher CTLA-4-expression than remaining patients and controls (figure 2C). Of note, because the number of borrelia-specific T cells was low in almost all patients and controls, further analysis of characteristic functional and phenotypical properties of borrelia-specific T cells was only possible for five controls and three patient samples, of which one patient also showed a particularly high CTLA-4-expression (figure 2C). CD27, a marker for T-cell differentiation, had similar expression levels on reactive T cells of controls and PFP-patients. However, while VZV-, HSV- and SEB-reactive T cells were mainly CD27-positive, CD27-expression on CMV-specific T cells was low (figure 2D), which is a known characteristic of CMV-specific T cells (28). The intranuclear expression of the proliferation marker Ki67 was generally low in pathogen-specific T cells of controls and PFP-patients. However, 8 out of 35 patients showed an increased percentage of Ki67-positive VZV-specific CD4 T cells (figure 2E). CTLA-4-, Ki67- and CD27-expression on SEB-reactive T cells did not show any significant differences between patients and controls.

**Figure 2:**
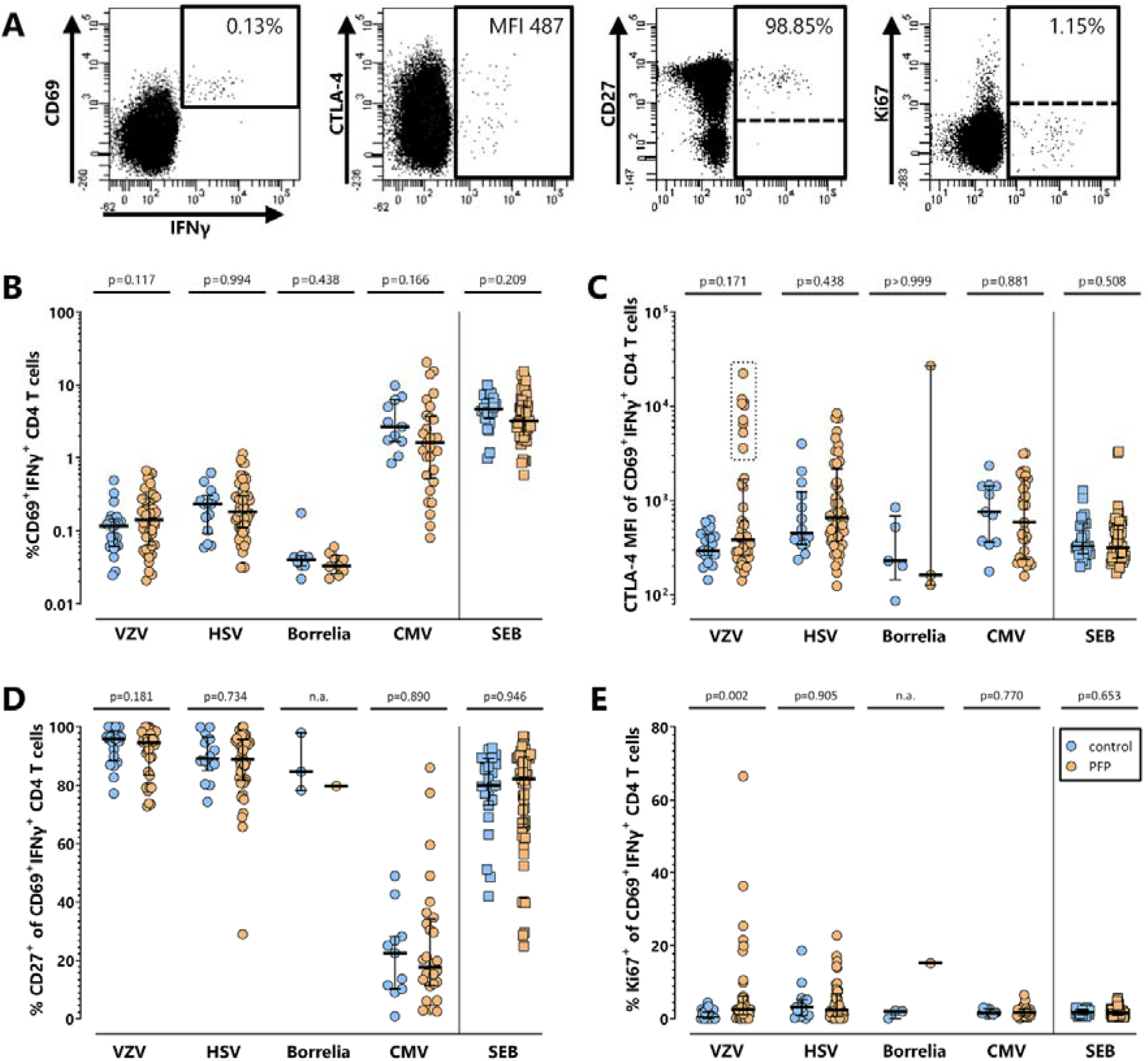
No distinct alterations in phenotype and proliferation of pathogen-specific T cells in patients with PFP. T-cell responses in controls and PFP-patients with detectable pathogen-specific T-cell frequencies were quantitatively and qualitatively characterized further. In **(A)** representative dot plots of VZV-stimulated CD4 T cells of a 28-years old PFP-patient are shown. Percentages of pathogen-specific CD4 T cells **(B)**, their expression of CTLA-4 **(C)**, and the percentage of pathogen-specific cells positive for CD27 **(D)** or Ki67 **(E)** were compared between controls (blue circles) and PFP-patients (orange circles). Numbers in panel A indicate percentage of reactive (CD69 IFNγ) among total CD4 cells and CTLA-4 expression (MFI) as well as percentages of CD27- or Ki67-positive CD4 T cells after stimulation with VZV-lysate. Bars in panel B-D represent median values. To ensure robust statistics, analysis in panel C-E was restricted to samples with at least 20 antigen-specific CD4 T cells. CMV, cytomegalovirus, CTLA-4, cytotoxic T-lymphocyte antigen 4; HSV, herpes-simplex viruses; IFN, interferon; MFI, median fluorescence intensity; PFP, peripheral facial palsy; SEB, *Staphylococcus aureus* enterotoxin B; VZV, varicella-zoster virus.

### Stable levels and phenotype of pathogen-specific T cells for at least two weeks

To assess potential dynamics in quantity and phenotypical properties of the pathogen-specific T cells early after the onset of PFP, the percentage and CTLA-4-, Ki67- and CD27-expression of pathogen-specific T cells of 36 PFP-patients was analyzed at first clinical presentation and 14 days thereafter (figure 3). Paired samples of 15 healthy individuals served as controls were no major changes in frequencies and expression patterns were expected. At the time of the second analysis, the median House-Brackmann grade of this subgroup of patients had not yet changed significantly (p=0.107). Hence, there were no significant differences in the pathogen-specific T-cell properties within this time frame, except for a slight decrease in CTLA-4-expression on VZV-specific CD4 T cells of PFP-patients. Although the decrease was significant (p=0.023), samples with high CTLA-4 expression at PFP-onset still had considerably high CTLA-4 expression levels two weeks later. Interestingly, samples of four patients with elevated percentages of Ki67^+^ VZV-specific CD4 T cells at PFP-onset showed a clear reduction of Ki67-positivity over time. Likewise, two other PFP-patients showed a similarly marked decrease in Ki67-positivity on HSV-specific T cells.

**Figure 3.**
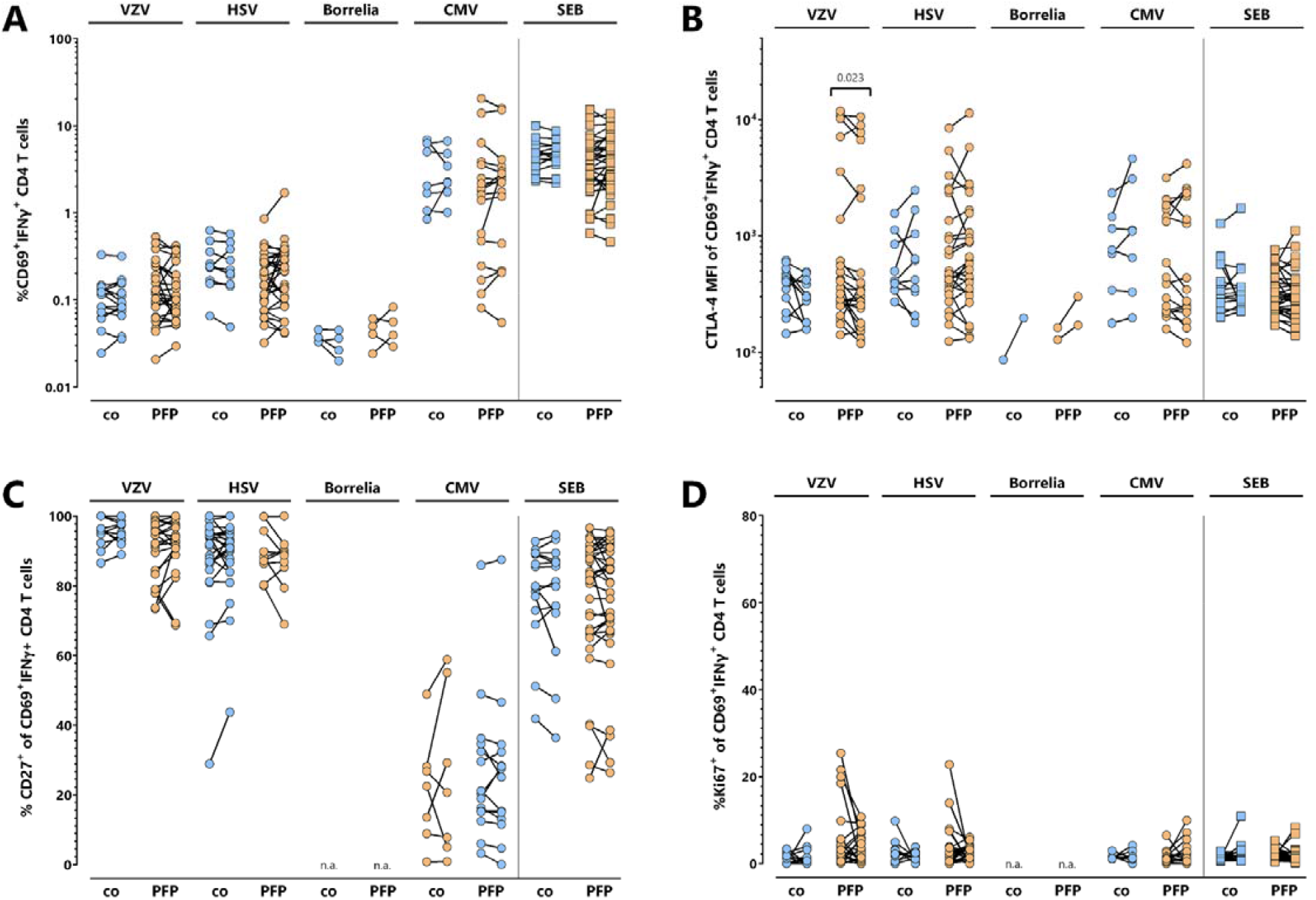
Stability of pathogen-specific T-cell characteristics within 14 days after clinical presentation. Pathogen-specific T-cell responses in 36 PFP-patients (orange circles) were analyzed at the beginning of symptoms and two weeks thereafter. In parallel, blood samples of 15 controls (blue circles) were additionally analyzed two weeks after the first sample recruitment. **(A)** Percentages of pathogen-specific and SEB-reactive CD4 T cells above detection limit were compared at both time points. **(B)** CTLA-4 expression as well as **(C)** CD27- and **(D)** Ki67-positivity of pathogen-specific T cells was compared at onset of symptoms. To ensure robust statistics, analysis of samples with positive pathogen-specific T-cell frequencies in panel B, C and D was restricted to samples with at least 20 antigen-specific CD4 T cells. CMV, cytomegalovirus, HSV, herpes-simplex viruses; IFN, interferon; PFP; peripheral facial palsy; VZV, varicella-zoster virus.

### High CTLA-4- and Ki67-expression of VZV-specific T cells as biomarker for VZV-related peripheral facial palsy

Based on available clinical parameters, two patients were diagnosed with Guillain-Barré syndrome. For the remaining 53 patients, a composite score was developed to classify PFP according to likelihood of an inflammatory or idiopathic cause, or as unclear (table 3). Based on this score, 8 out of 53 PFP-patients showed predominant signs of an inflammatory cause with or without detectable pathogen, whereas 28 PFP-patients were classified as idiopathic. The diagnosis of PFP was classified as unclear in 17 patients (figure 4A).

**Table 3:**
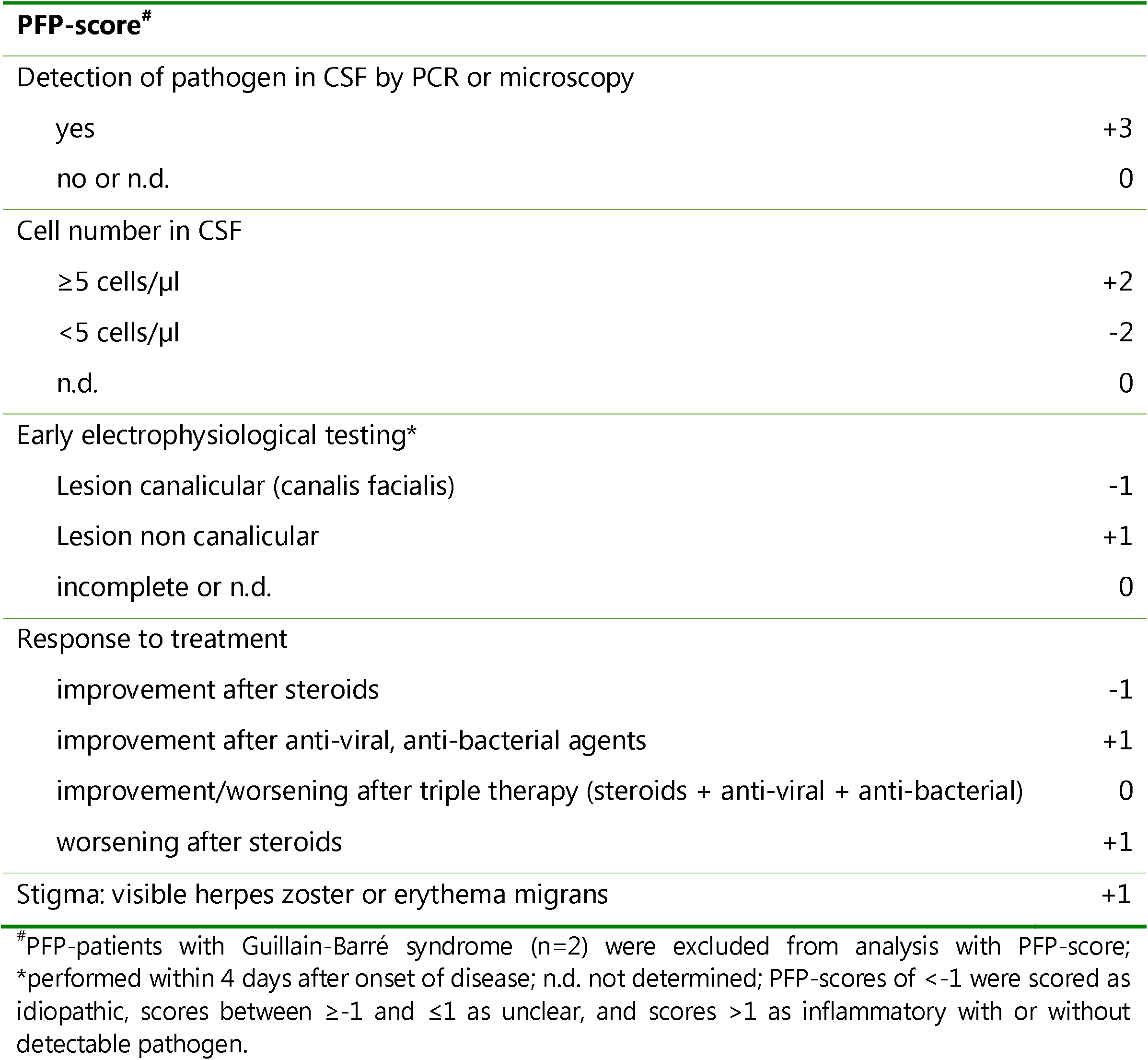
Clinical parameters for calculation of the PFP-score.

| **PFP-score<sup>#</sup>** |  |
| --- | --- |
| Detection of pathogen in CSF by PCR or microscopy |  |
| yes | +3 |
| no or n.d. | 0 |
| Cell number in CSF |  |
| ≥5 cells/μl | +2 |
| <5 cells/μl | -2 |
| n.d. | 0 |
| Early electrophysiological testing* |  |
| Lesion canalicular (canalis facialis) | -1 |
| Lesion non canalicular | +1 |
| incomplete or n.d. | 0 |
| Response to treatment |  |
| improvement after steroids | -1 |
| improvement after anti-viral, anti-bacterial agents | +1 |
| improvement/worsening after triple therapy (steroids + anti-viral + anti-bacterial) | 0 |
| worsening after steroids | +1 |
| Stigma: visible herpes zoster or erythema migrans | +1 |

<sup>#</sup>PFP-patients with Guillain-Barré syndrome (n=2) were excluded from analysis with PFP-score;
\*performed within 4 days after onset of disease; n.d. not determined; PFP-scores of <-1 were scored as idiopathic, scores between ≥-1 and ≤1 as unclear, and scores >1 as inflammatory with or without detectable pathogen.

**Figure 4:**
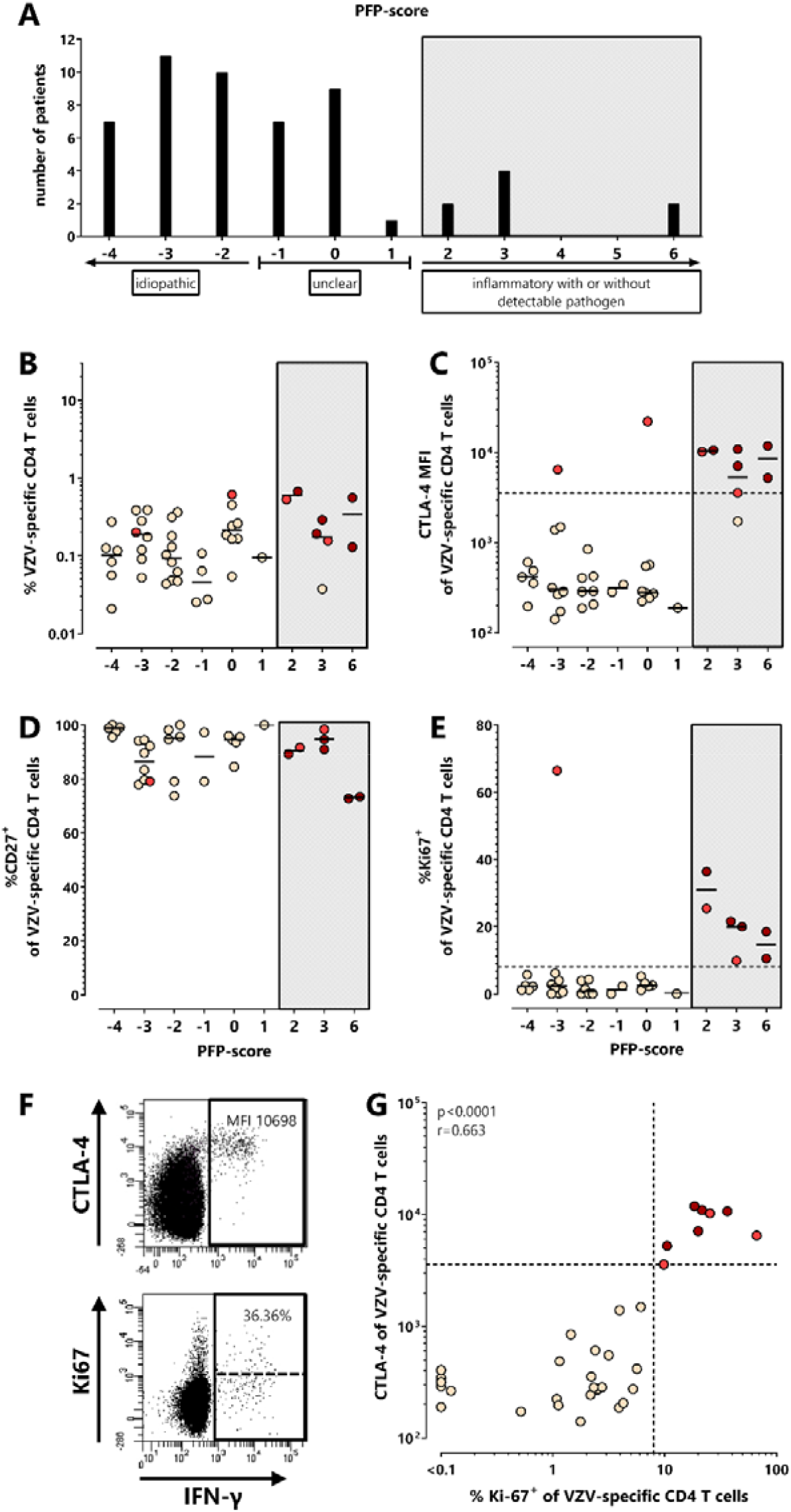
Characteristic alterations in CTLA-4 and Ki67-expression of VZV-specific T cells in patients with VZV-associated PFP. **(A)** Patients with PFP were subclassified according to a composite PFP-score based on the results of available routine clinical diagnostics (see table 3, n=53, 2 patients with Guillain-Barré syndrome were not included) to allow determination of the most probable cause of PFP. Scores <-1 were scored as idiopathic (n=28), scores between ≥-1 and ≤1 as unclear (n=17), and scores >1 as inflammatory with or without detectable pathogen (n=8, including 5 PFP-patients with VZV-induced skin disease (3x zoster oticus, 1x Ramsay Hunt zoster, 1x with concomitant cervical (C2) zoster efflorescence)). **(B)** VZV-specific CD4 T-cell levels of PFP-patients with positive VZV T-cell status (n=45) were compared depending on the PFP-score. Expression of **(C)** CTLA-4 (n=39), **(D)** CD27 (n=34), and **(E)** Ki67positivity (n=34) of VZV-specific T cells is shown in patients with different PFP-scores. **(F)** Dot plots of a 79-years old PFP-patient with VZV-reactivation show characteristic expression of CTLA-4 (upper panel) and Ki67 (lower panel) on (IFNγ+) CD4 T cells after in-vitro stimulation with VZV-antigen. **(G)** CTLA-4 and Ki67-expression of VZV-specific CD4 T cells was correlated among all VZV T-cell positive samples of PFP-patients where both markers were available (n=34). To ensure robust statistics, analysis in panel C, D, E and G was restricted to samples with at least 20 antigen-specific CD4 T cells. Dotted lines in panel C, E and G represent cut-offs (3579 MFI for CTLA-4 (26), and 8% for %Ki67-positive) which discriminated best between VZV-associated PFPs and non VZV-associated PFPs. Patients with dark red and light red symbols refer to patients with high CTLA-4 expression levels on VZV-specific T cells and/or high percentage of Ki67-positive VZV-specific T cells. One patient (light red) did not have sufficient blood for CD27- and Ki67-staining. Numbers in panel F indicate CTLA-4-MFI and percentage of Ki67-positive of VZV-reactive CD4 T cells. CTLA-4, cytotoxic T-lymphocyte antigen 4; IFN, interferon; MFI, median fluorescence intensity; PFP, peripheral facial palsy; VZV, varicella-zoster virus.

Among the 8 patients with inflammatory PFP-score, 5 had VZV-related skin disease with a score ≥2 (3 patients with zoster oticus, 1 patient with Ramsay hunt zoster and 1 patient with concomitant cervical (C2) zoster efflorescence). All patients with detectable VZV-specific T cells are shown in figure 4B-G. While the percentage and CD27-expression of VZV-specific CD4 T cells of the 5 patients with zoster manifestations (labelled in dark red) did not differ from those of patients with lower PFP-scores (figure 4B and D), CTLA-4- and Ki67-expression were considerably higher (figure 4C and E), with typical dot plots shown in figure 4F. Their CTLA-4-MFI was above the cut-off previously established for active VZV-infection (26). Interestingly, a similar VZV-specific CD4 T-cell phenotype (CTLA-4-MFI >3579 and >8% Ki67-expressing cells) was also found in four more PFP-patients (labelled in light red), although the causative pathogen was either not detectable (n=2, score 2 and 3), standard clinical data were unavailable (n=1, score 0) or PFP in one pregnant woman was even classified as “idiopathic” (n=1, score -3). Among those, direct and indirect VZV-diagnostic was only performed in 2 out of 4 patients. While VZV-DNA was not detectable, these two patients (with score 2 and 3) had an intermediate or positive VZV-ASI. Overall, expression levels of CTLA-4 and Ki67 on VZV-specific T cells showed a significant correlation (r=0.663, p<0.0001, figure 4G). Of note, in patients with altered phenotype of VZV-specific T cells, the properties of HSV-, CMV- and SEB-reactive T cells were unaltered and similar as in patients with normal VZV-specific T-cell profile (figure S1), which emphasizes that alterations in phenotype were VZV-specific and did not affect effector T cells in general.

### Altered phenotype of borrelia-specific T cells in a case with neuroborreliosis

In general, frequencies of borrelia-specific T cells were low or undetectable in both seronegative and seropositive individuals, and did not show any evidence for phenotypical alterations. An exception was a 9-years old boy with acute PFP and recent history of untreated erythema migrans (1 month before) who had 0.04% of borrelia-specific T cells. Interestingly, despite this low percentage, these cells showed particularly high CTLA-4- and Ki67-expression, whereas SEB-reactive T-cell properties were unaltered except for a slightly elevated CTLA-4 expression (figure S2). Likewise, VZV- and CMV-specific T cells had a normal phenotype and HSV-specific T cells were below detection limit. Of note, this was the only patient with both positive borrelia-specific IgG and IgM (table 2).

## Discussion

In this study, we performed a detailed quantitative and functional analysis of immune responses against pathogens such as VZV, borrelia and HSV which are associated with peripheral facial palsy (4-7, 29). In general, we found high concordance of virus-specific antibody and T-cell responses in our study cohort. While most tested parameters did not differ between patients and controls, patients had a significantly higher HSV-seroprevalence, which may support a causative role of HSV in the etiology of PFP. Nevertheless, HSV-specific T-cell levels or their phenotypical characteristics did not differ in patients and controls. In contrast, we found a distinct fraction of PFP-patients with a strong expression of CTLA-4 and Ki67 on their VZV-specific T cells. This immunological alteration may hold promise as a biomarker for a causal role of VZV among individuals with PFP of unknown cause.

We have previously shown that CTLA-4 expression is specifically upregulated on VZV-specific T cells in individuals with active herpes zoster (25) or patients with VZV-related diseases of the central nervous system such as meningitis or encephalitis with a 100% sensitivity and 100% specificity for individuals with CTLA-4 MFI above the threshold of 3579 (26). This previous study also identified 3 more patients with high CTLA-4 expression levels with unclear CNS-diagnosis, but a diagnosis of PFP (26). The PFP-patients in the present study were heterogeneous regarding their clinical diagnosis of the underlying cause of PFP. As a consequence, the whole cohorts of controls and PFP-patients did not show any differences in VZV-specific T-cell characteristics. However, when applying the threshold previously established for patients with acute VZV-induced CNS-disease (26), we identified 9 patients where CTLA-4 expression of VZV-specific CD4 T cells was strongly increased. This phenotype was specific for VZV-specific T cells, as T cell characteristics with specificity towards other pathogens were similar as in controls. Among these patients, five had acute symptomatic VZV-reactivation with typical skin disease manifestations and two additional patients showed elevated VZV-ASI, suggesting VZV-reactivation (zoster sine herpete), whereas a potential VZV-involvement or alternative diagnosis remained unclear in the other 2 patients based on clinical anamneses and standard laboratory diagnostics. It is therefore tempting to speculate whether these alterations may indicate a potential involvement of VZV, which may be tested in future studies by empirical treatment with appropriate antiviral therapeutics. Notably, in addition to increased CTLA-4 levels, a large fraction of VZV-specific T cells of these 9 patients was also Ki67-positive (9.8-66.4%), indicating recent proliferation. Based on a similar observation in patients with active tuberculosis who showed higher percentages of Ki67-positive cells among *Mycobacterium tuberculosis-*specific T cells than latently infected individuals (30), increased percentages of Ki67-positive among VZV-specific T cells may also argue for a causative role of VZV. Interestingly, one of these before mentioned 4 patients was VZV-IgG seronegative at initial measurement. Thus, unlike VZV-reactivations, PFP in this patient may have been triggered by a recent history of symptomatically untypical primary infection.

Follow-up data two weeks after the first sample acquisition revealed that the percentage of VZV-specific CD4 T cells and their CTLA-4- and CD27-expression remained almost stable or were only slightly altered. In contrast, Ki67-protein seems to be degraded during this time frame as indicated by four patients with increased percentage of Ki67-positive VZV-specific cells at time of PFP-symptoms and lower frequencies 14 days thereafter. As PFP-patients with increased CTLA-4 expression of VZV-specific T cells had a similar cellular phenotype two weeks later, this biomarker may be useful to identify VZV-associated PFPs during a diagnostic time frame of up to 14 days after onset of symptoms. Despite the fact that we have not performed any analyses at later time points in the present study, we have previously shown that characteristic phenotypical (including CTLA-4 expression) and functional alterations of VZV-specific CD4 T cells in patients with acute herpes zoster reverted back to values almost comparable to those of non-symptomatic individuals 3 months post VZV-reactivation (25). Thus, future studies should investigate dynamic changes in VZV-specific T-cell characteristics to address whether the properties of VZV-specific T-cell responses in PFP-patients will also normalize after a longer period of time.

Interestingly, HSV-seroprevalence in our PFP-patient group was significantly higher (90.9%) than in our control group which may indicate a potential causative role of HSV in some patients with PFP. Although an HSV-seroprevalence of 52% in our control group may be slightly underrepresented, the high seroprevalence among PFP-patients is remarkable, as HSV-1-prevalence in the general population in Germany has rather been decreasing from 82.1% in 1997–1999 to 78.4% in 2008-2011 (31). If the analysis is restricted to PFP-patients scored as “unclear” or “idiopathic” using standard diagnostic parameters, HSV-seropositivity was even above 95% (43/45) and thus seems to be higher than in the general German population and therefore may further indicate involvement of HSV in PFP. Support for a potential HSV-involvement is given by observations from a small study showing that HSV-DNA was detectable in 79% of facial nerve endoneural fluid and posterior auricular muscle of patients with “idiopathic” PFP, whereas there was no HSV-DNA in VZV-triggered PFPs or controls (3). However, based on the small sample size of that study and conflicting results of larger studies on the clinical benefit of aciclovir treatment of patients with Bell’s palsy, further studies are needed to further substantiate HSV as a causative virus in PFP (2, 4, 16, 32-36). Furthermore, apart from increased HSV-seroprevalence in our population of PFP-patients, our study did not show any characteristic alterations in the phenotype and function of HSV-specific T-cells, which may be due to the fact that HSV-reactivations may rather be associated with local recruitment of specific T cells without measurable systemic changes. Thus, unlike VZV-specific T cells, HSV-specific T-cell analysis in circulation is unlikely to be useful to individually identify patients whose PFP-symptoms may be HSV-triggered.

Our study is limited by the in part low number of pathogen-specific T cells, especially after borrelia-specific stimulations. Thus, the presence of sufficient numbers of pathogen-specific T cells seems critical to enable further characterization of CTLA-4- and/or Ki67-expression. However, despite the generally low percentage of borrelia-specific T cells in our cohorts, the example of a 9-years old boy with confirmed neuroborreliosis demonstrates that borrelia-specific T cells might be detectable during active disease thus enabling further characterization of phenotypical markers associated with altered T-cell function. Thus, as with VZV, upregulation of CTLA-4 and Ki67 on borrelia-specific T cells may also be indicative of a specific involvement of borrelia, which warrants further study. As a further limitation, the cause of PFP remains unclear in a large fraction of patients. Therefore, a causal association of altered T-cell characteristics with VZV-involvement remains speculative in the patients with unclear diagnosis. In this regard, interventional studies assessing response to treatment with either specific anti-viral or anti-bacterial agents may provide evidence as to whether high levels of CTLA-4 on VZV- or borrelia-specific CD4 T cells may allow identification of individuals with VZV- or borrelia-induced PFP.

In conclusion, although the quantity and phenotype of VZV-specific T-cell responses did not generally differ between PFP-patients and healthy controls, the distinct increase in CTLA-4- and Ki67-expression of VZV-specific cells allows for identification of VZV-related PFP. This may even extend to patients where conventional clinical and laboratory diagnosis is unclear or limited. Combined with first evidence from a patient with neuroborreliosis, analysis of pathogen-specific T-cell properties may be suitable to identify VZV or borrelia as infectious causes in PFP, whereas HSV-specific T cells do not seem to be useful to identify involvement of HSV. However, the high HSV-seroprevalence in patients with idiopathic PFP is a hint for an underestimated role of HSV during development of PFP. Our findings need to be confirmed by larger sample size and an interventional study with specific anti-viral or antibiotic therapy; in this case, determination of altered phenotype of pathogen-specific T cells could be applied as an adjunct tool towards improved diagnostics of PFP.

## Materials and methods

### Recruitment of the study population

Patients with clinically diagnosed acute PFP were recruited at the Department of Neurology, Department of Otorhinolaryngology and Department of Pediatrics and Neonatology of Saarland University between September 2017 and February 2019. Twenty-three age-matched healthy individuals served as control group. Clinical grading of PFP was determined using the House-Brackmann score (1). All patients received standard routine diagnostics including direct detection of pathogens in cerebrospinal fluid (CSF) by PCR or microscopy, determination of cell number in CSF, electrodiagnostic testing, assessment of response to treatment, and/or screening for VZV- or borrelia-related skin manifestation. Diagnostic parameters and treatment were chosen by the treating physicians and decisions were unaffected by results of the current study. To address clinical grading and stability of immunological parameters during follow up, a subgroup of PFP-patients and controls were analyzed a second time approximately two weeks after the first sample acquisition. The study was approved by the local ethics committee (Ärztekammer des Saarlandes, reference number 158/13) and all participants or parents of participating children gave written informed consent.

### Quantitative and functional analysis of pathogen-specific T cells

Analysis of pathogen-specific T-cell responses in whole blood samples was performed as described before (25, 37). In brief, heparinized whole blood was stimulated with VZV-lysate, HSV-1/-2-lysate, CMV-lysate (each 32μl/ml; Virion/Serion) and a mix of bulk antigens from *borrelia garinii* and *borrelia afzelii* (each 10μg/ml; Virion/Serion), respectively. Stimulations with uninfected control lysates (32μl/ml; Virion/Serion) and 2.5μg/ml *Staphylococcus aureus* enterotoxin B (SEB; Sigma) served as negative and positive control, respectively. All stimulations were carried out in the presence of 1μg/ml anti-CD28 and anti-CD49d costimulatory antibodies (BD Biosciences). After 2h, brefeldin A was added for intracellular accumulation of induced cytokines. After additional 4h, cells were EDTA-treated and fixed.

Two separate staining reactions were performed to analyze surface markers (CD4, CD27, CD69, CTLA-4) and intracellular molecules (IFN*γ*, Ki67) after permeabilization of cells with a saponin-containing buffer (BD Biosciences)). Ki67, a marker for recent proliferation was intranuclearly detected using the “Foxp3/Transcription Factor Staining Buffer Set” (ThermoFisher) in combination with anti-Ki67 antibodies (BD Biosciences). 300μl of whole blood were used per stimulatory reaction and staining. If blood volume was limited, staining for CD27 and Ki67 was omitted. At least 10000 CD4 T cells per sample were flow-cytometrically analyzed using a BD FACS Canto II and the BD FACSDiva software version 6.1.3.

### Semi-quantitative and quantitative analysis of pathogen-specific antibodies

VZV-, HSV- and CMV-specific IgG-levels in blood were quantified using anti-IgG enzyme-linked immunosorbent assays (ELISA, Euroimmun, Lübeck, Germany). Ratios of VZV- and HSV-specific IgAs and HSV-specific IgMs were semi-quantitatively determined by anti-IgA and -IgM ELISA (Euroimmun), respectively. Borrelia-specific IgGs and IgMs were screened using the chemiluminescence immunoassay (CLIA) technology (Liaison®, DiaSorin). Intermediate and positive screening results were subsequently confirmed by line-immunoblot (Borrelia ViraStripe®, Viramed). Cut-offs to define negative, intermediate and positive responses were defined as per manufacturer’s instructions.

### PFP-score as correlate of clinical standard parameters

PFP-patients were subclassified according to results of available routine clinical diagnostics. These results were compiled in a composite PFP-score as detailed in table 3 to allow assignment of the most probable cause of PFP. Scores <-1 were scored as idiopathic, scores between ≥-1 and ≤1 as unclear, and scores >1 as inflammatory with or without detectable pathogen.

### Statistical analysis

Statistical analysis was performed using GraphPad-Prism 10.0.0. Non-normally distributed variables between controls and PFP-patients were compared using Mann-Whitney test. Analysis of parametric values (age, time between 1^st^ and 2^nd^ blood sampling) was performed using unpaired t-test. Wilcoxon matched pairs tests were performed to compare T-cell responses at the two time points of one individual. Differences in sex distribution or serostatus between groups were analyzed by Fisher’s exact test. Cut-off values for VZV-, HSV- and CMV-specific CD4 T cells (0.02%, 0.025% and 0.05%, respectively) were calculated by ROC-analysis of T-cell responses in seronegative and seropositive healthy individuals and/or as defined before (25, 38). As borrelia-specific antibody responses are of limited value, calculation of a cut-off for borrelia-specific T cells based on serology was not considered reasonable. Therefore, the lowest cut-off of 0.02% was chosen for borrelia-specific T cells in the context of this study.

## Supporting information

Mohammad_supplement

## Data Availability

All data produced in the present study are available upon reasonable request to the authors

## Abbreviations

ASI: antibody specific index
CMV: cytomegalovirus
CTLA-4: cytotoxic T-lymphocyte antigen-4
HSV: herpes-simplex virus
IFN: interferon
MFI: median fluorescence intensity
PFP: peripheral facial palsy
SEB: Staphylococcus aureus enterotoxin B
VZV: varicella-zoster virus

## Acknowledgements

The authors thank Candida Guckelmus and Lisa Lieblang for excellent technical assistance.

## Acknowledgements

The authors thank all participants to this study. Expert technical assistance by Candida Guckelmus is acknowledged. Financial support was provided in part by a intramural research grant by HOMFOR to D.S..

## Author contributions

D.S., M.F., T.S. and M.S. designed the study; D.S., M.F., T.S. and M.S. designed the experiments, L.M., D.S., and T.S. performed experiments; M.L., M.F., G.W., M. F.-B., K.F., U.D., B.S., M.Z., B.C.G., U.S. contributed to study design, patient recruitment, and clinical data acquisition. M.L., M.F., D.S., U.S., T.S. and M.S. performed statistical analysis. M.F., D.S., T.S., and M.S. supervised all parts of the study, and performed analyses; D.S., M.F., and M.S. wrote the manuscript. All authors approved the final version of the manuscript.

## Supporting documents

**Figure S1:**
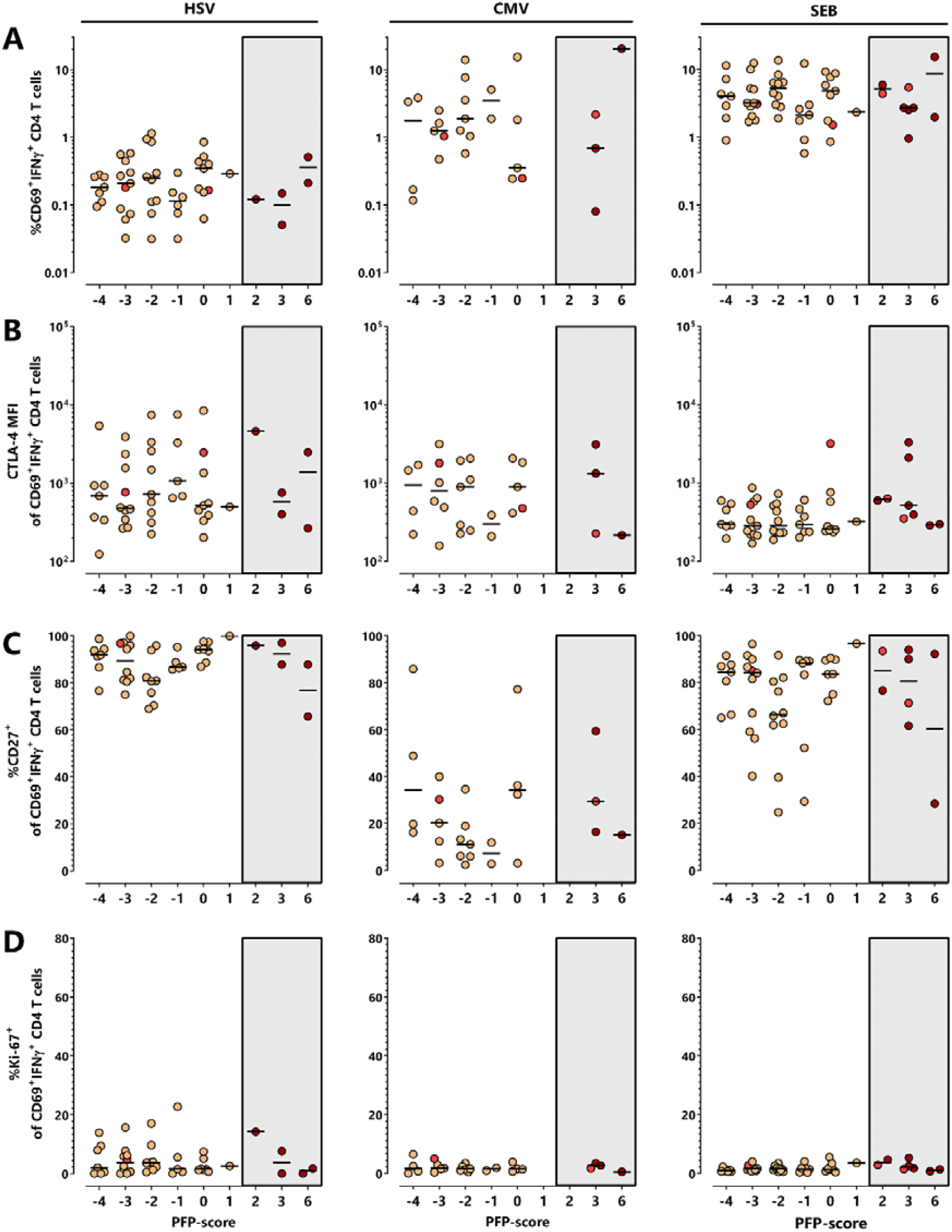
Distribution of the percentage and phenotype of HSV-, CMV- and SEB-reactive CD4 T cells according to PFP-score. PFP-patients (n=53) were subclassified according to PFP-score (s. table 3 and figure 4A). Scores <-1 were scored as idiopathic, scores between ≥-1 and ≤1 as unclear, and scores >1 as inflammatory with or without detectable pathogen. **(A)** HSV-, CMV- and SEB-reactive CD4 T-cell levels of T-cell positive individuals were compared between PFP-patients with different PFP-score. In addition, CTLA-4 **(B)**, CD27-**(C)** and Ki67-expression **(D)** of reactive T cells was analyzed with regard to PFP-score. To ensure robust statistics, analysis in B, C and D was restricted to samples with at least 20 antigen-specific CD4 T cells. Lines represent median values. Patients with dark red and light red symbols refer to patients with high CTLA-4 expression levels on VZV-specific T cells and/or high percentage of Ki67-positive VZV-specific T cells (see figure 4). CMV, cytomegalovirus; CTLA-4, cytotoxic T-lymphocyte antigen 4; IFN, interferon; HSV, herpes-simplex viruses; MFI, median fluorescence intensity; PFP, peripheral facial palsy; SEB, *Staphylococcus aureus* enterotoxin B.

**Figure S2:**
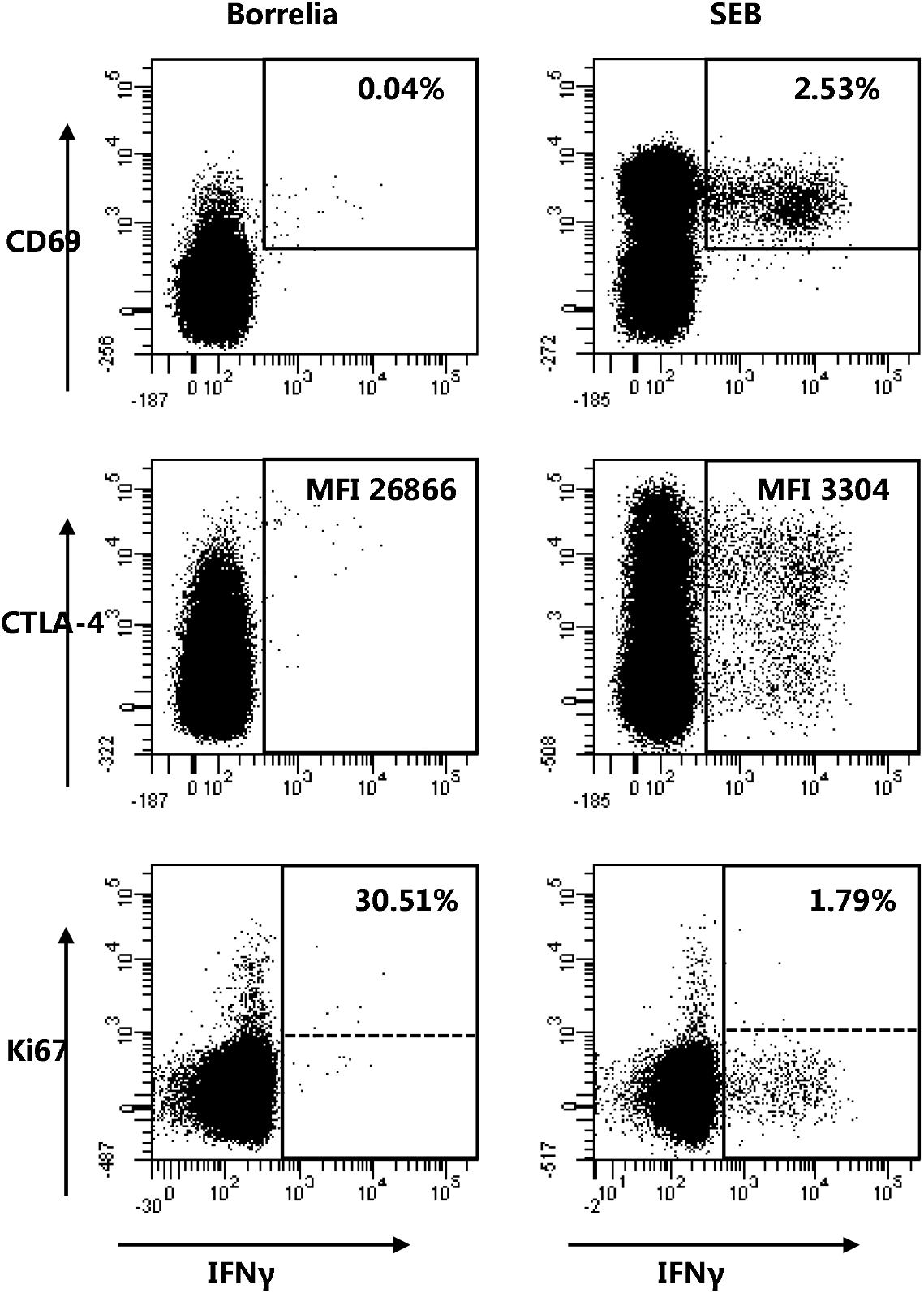
Distinct changes of borrelia-specific T-cell properties in a case of neuroborreliosis-related PFP. Borrelia-specific **(A)** and SEB-reactive **(B)** CD4 T cells of a 9-years old boy with acute PFP and confirmed neuroborreliosis were determined after antigen-specific stimulation and flow cytometric detection. Numbers in each dot plot indicate percentages of reactive (CD69 IFNγ) CD4 T cells (upper panels), CTLA-4 (middle panels) and Ki67-expression of reactive CD4 T cells (lower panels), respectively. Follow-up data of this patient were not available. CTLA-4, cytotoxic T-lymphocyte antigen 4; IFN, interferon; MFI, median fluorescence intensity; PFP, peripheral facial palsy; SEB, *Staphylococcus aureus* enterotoxin B.

