## Supplementary material for "Alterations in pathogen-specific cellular and humoral immunity associated with acute peripheral facial palsy of infectious origin": Mohammad_supplement

### Supporting documents

#### Figure S1

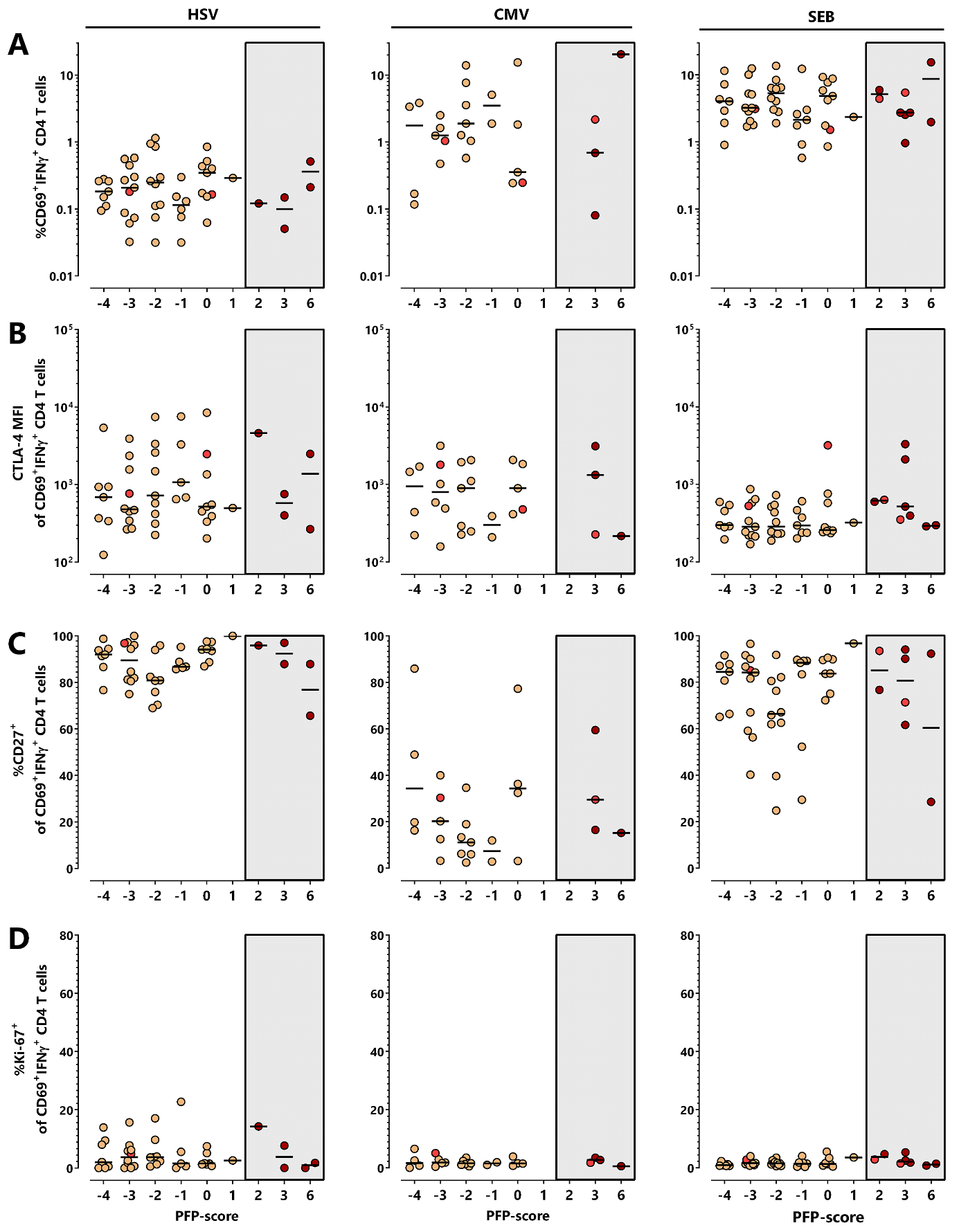

**Figure S1: Distribution of the percentage and phenotype of HSV-, CMV- and SEB-reactive CD4 T cells according to PFP-score.** PFP-patients (n=53) were subclassified according to PFP-score (s. table 3 and figure 4A). Scores <-1 were scored as idiopathic, scores between ≥-1 and ≤1 as unclear, and scores >1 as inflammatory with or without detectable pathogen. **(A)** HSV-, CMV- and SEB-reactive CD4 T-cell levels of T-cell positive individuals were compared between PFP-patients with different PFP-score. In addition, CTLA-4 **(B)**, CD27- **(C)** and Ki67-expression **(D)** of reactive T cells was analyzed with regard to PFP-score. To ensure robust statistics, analysis in B, C and D was restricted to samples with at least 20 antigen-specific CD4 T cells. Lines represent median values. Patients with dark red and light red symbols refer to patients with high CTLA-4 expression levels on VZV-specific T cells and/or high percentage of Ki67-positive VZV-specific T cells (see figure 4). CMV, cytomegalovirus; CTLA-4, cytotoxic T-lymphocyte antigen 4; IFN, interferon; HSV, herpes-simplex viruses; MFI, median fluorescence intensity; PFP, peripheral facial palsy; SEB, *Staphylococcus aureus* enterotoxin B.

#### Figure S2

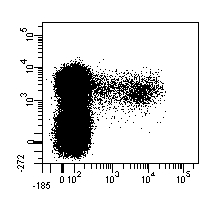

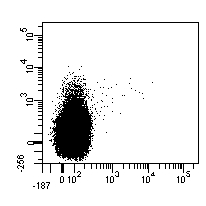

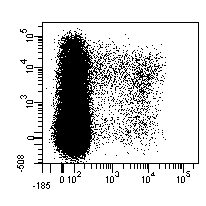

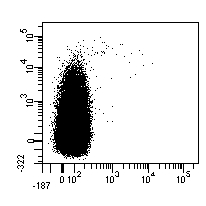

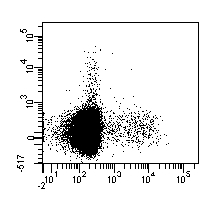

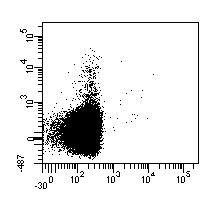

**30.51%**

**1.79%**

**MFI 26866**

**MFI 3304**

**0.04%**

**2.53%**

**Borrelia**

**SEB**

**IFNγ**

**Ki67**

**IFNγ**

**CTLA-4**

**CD69**

**Figure S2: Distinct changes of borrelia-specific T-cell properties in a case of neuroborreliosis-related PFP.** Borrelia-specific **(A)** and SEB-reactive **(B)** CD4 T cells of a 9-years old boy with acute PFP and confirmed neuroborreliosis were determined after antigen-specific stimulation and flow cytometric detection. Numbers in each dot plot indicate percentages of reactive (CD69^+^IFNγ^+^) CD4 T cells (upper panels), CTLA-4 (middle panels) and Ki67-expression of reactive CD4 T cells (lower panels), respectively. Follow-up data of this patient were not available. CTLA-4, cytotoxic T-lymphocyte antigen 4; IFN, interferon; MFI, median fluorescence intensity; PFP, peripheral facial palsy; SEB, *Staphylococcus aureus* enterotoxin B.
